# Why Risk Factors Minimally Change the ROC Curve AUC

**DOI:** 10.1101/2022.02.24.22271481

**Authors:** Ralph H. Stern

## Abstract

Novel risk factors that improve statistical measures of fit on addition to established clinical prediction models often minimally change measures of discrimination (ROC curve AUC or c-index). As a result, measures of discrimination have been suggested to be insensitive in the evaluation of such models.

To understand this phenomenon, it is necessary to focus on the population risk distributions produced by models with and without the risk factor. This is because these risk distribution fully determines the risk distributions of cases/patients and controls/nonpatients, which in turn fully determine the ROC curve and its AUC. Broader population risk distributions result in larger ROC curve AUCs.

A fully independent risk factor with a relative risk of 2 added to the standard cardiovascular risk model produces risk distributions of those with or without the risk that are clearly different (which is evaluated by statistical measures of fit), while minimally broadening the population risk distributions (which is evaluated by measures of discrimination). The reason for this is that although addition of the risk factor replaces every risk stratum with higher and lower risk strata, this depopulated risk stratum is largely repopulated by similar splitting in neighboring risk strata. The interweaving of the the up and down migration paths to and from every point on the risk distribution results in a largely compensatory shuffling of risk assignments with minimal changes in the ROC curve AUC.

---

Basic research identifies novel disease risk factors every year. In the finance literature, the large number of factors influencing investment returns has been called “a zoo of new factors”. (1) Since preventive measures are often allocated based on risk, evaluating members of the disease risk factor zoo for clinical adoption is a major challenge. A recurrent observation is that new risk factors may improve statistical measures of fit when added to multivariate models, but not measures of discrimination (the ROC curve AUC or c-index). This has led to the suggestion that measures of discrimination are insensitive in evaluating risk prediction models and that they are being abused for this purpose. (2,3)

As an alternative, analysis of reclassification has been suggested to address clinical utility. (2) Here predicted probabilities are grouped into risk categories used in guidelines and the changes in risk category assignment for individuals on addition of the new risk factor are evaluated. Based on this suggestion, new statistical measures were developed. (4)

The prior literature on these topics has focused on statistics and not provided an explanation for why risk factors minimally change the ROC curve AUC. Here a graphical approach is adopted to provide insight into these issues.

## Methods

The pooled cohort equations (PCE) based on the Framingham risk factors are used clinically to risk stratify the population by 10-year risk of fatal or nonfatal myocardial infarction or stroke. A beta distribution approximating the risk distribution resulting from application of the PCE to the US adult population was derived strictly to illustrate underlying principles. This was used as a basis for calculating the results of addition of a risk factor. Mathematica was used to perform calculations and prepare graphs.

### PCE risk distribution

Based on application of the PCE to the NHANES study population, the breakdown of the US adult population into risk categories was reported as: <0.05, 0.581; 0.05 to 0.10, 0.220; and >=10, 0.252. (5) A beta distribution with alpha of 0.749751 and beta of 9.86786 matches these categories.

### Distributions of low risk and high risk

The effects of adding an independent risk factor with a relative risk (RR) and a prevalence (PREV) can be calculated. This splits a risk stratum into a low-risk and high-risk stratum or a population into a low-risk and high-risk subgroup. If the original risk was x, the low risk stratum/subgroup would have a risk of x/(1+PREV(RR-1)), while the high risk stratum/subgroup would have a risk of RR*x/(1+PREV(RR-1)), since (1-PREV)*x/(1+PREV(RR-1))+PREV*RR*x/(1+PREV(RR-1))=x. For a risk factor with a relative risk of 2 and a prevalence of 0.25, the low-risk subgroup/stratum has a risk of x/1.25 and the high-risk subgroup/stratum has a risk of 2*x/1.25.

Since the PCE mean risk is 0.0706139, the low risk subgroup (without the risk factor), constituting 0.75 of adults, would have a mean risk of 0.0706139/1.25 or 0.0564911 and the high risk subgroup (with the risk factor), constituting 0.25 of adults, would have a mean risk of 2*0.0706139/1.25 or 0.112982. The independent variable in the population beta risk distribution is multiplied by 1.25 to obtain the low-risk beta distribution. In addition, the densities need to be multiplied by 1.25 to produce a probability density function. This ensures the area under the curve is 1 and the distribution has the correct mean. Similarly, the independent variable in the population beta risk distribution is multiplied by 0.625 to obtain the high-risk beta distribution and the densities need to be multiplied by 0.625. The area under the curve from 0 to 1 for the high-risk beta distribution, 0.999968, is not quite 1 as a small tail of this distribution exceeds a risk of 1.

### PCE+RF risk distribution

The PCE+RF population risk distribution is simply the weighted sum of the low-risk and high risk beta distributions. A very small fraction (0.000008) of this distribution exceeds a risk of 1 as 0.000032 of the high-risk distribution exceeds a risk of 1

### Calculation of density of 0.05 PCE+RF risk stratum from donor PCE strata

When a risk factor is added to the PCE, 0.75 of the 0.05 risk stratum migrates to a lower risk stratum in the PCE+RF distribution and 0.25 to a higher risk stratum. The 0.05 risk stratum in the PCE+RF distribution is populated by receipt of 0.75 of the 0.0625 PCE risk stratum and 0.25 of the 0.03125 risk stratum. However, for the reasons discussed above in calculating the low-risk and high-risk distributions, the densities need to be multiplied by the same adjustment factors, 1.25 and 0.625.

### Reclassification

A statin treatment criterion of >0.075 10-year risk is used to calculate reclassification on addition of the risk factor to the PCE.

Individuals without the risk factor whose PCE risk is between 0.075 and 0.09375 will be reclassified to a PCE+RF risk below 0.075. This is because 0.09375/1.25 is 0.075. This would apply to 0.75 of the individuals in this risk range.

Individuals with the risk factor whose PCE is between 0.046875 and 0.075 will be reclassified to a PCE+RF risk above 0.075. This is because 0.046875*2/1.25 is 0.075. This would apply to 0.25 of the individuals in this risk range.

## Results

The risk distributions and cumulative risk distributions based on the PCE for the overall population, the low-risk subgroup without the risk factor, and high-risk subgroup with the risk factor, shown in figure 1, are strikingly different. However, the risk distributions for the overall population from the PCE and from the PCE+RF, shown in figure 2, are almost superimposable.

**Figure 1.**
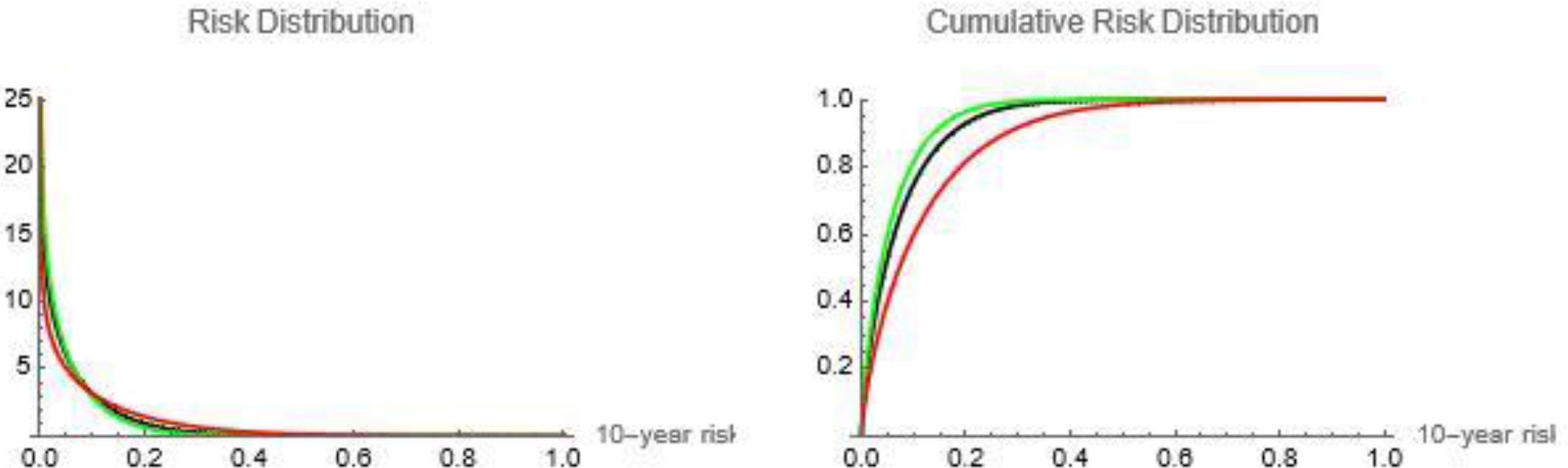
Risk distributions and cumulative risk distributions of PCE, low risk subgroup, and high risk subgroup. PCE: black; low risk: green; high risk: red

**Figure 2.**
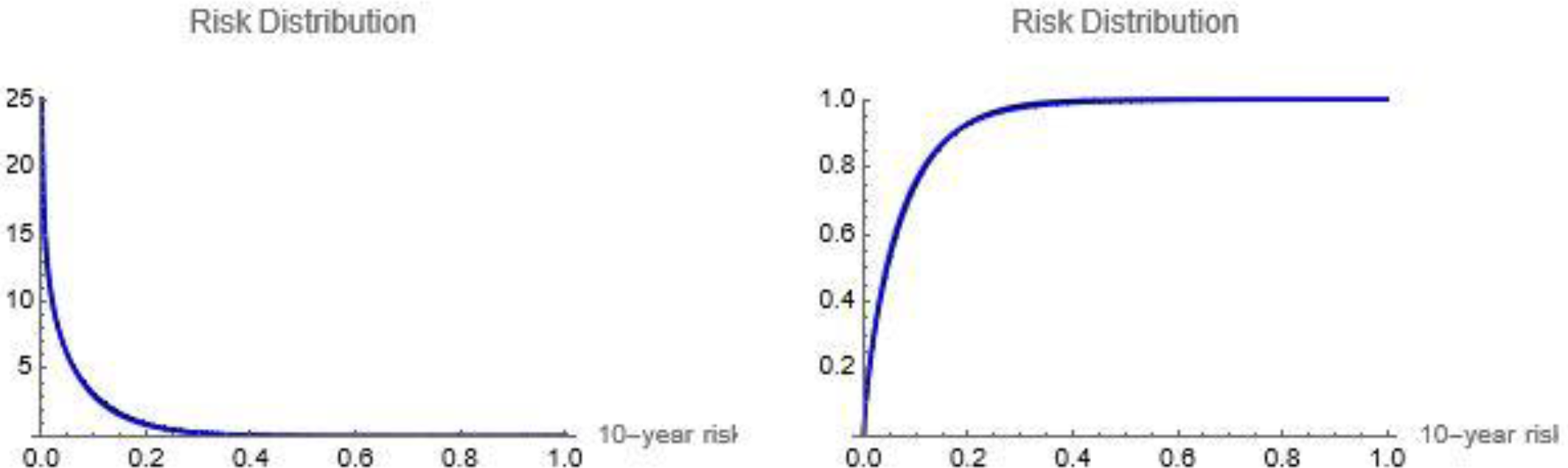
Risk distributions and cumulative risk distributions of PCE and PCE+RF PCE: black; PCE+RF: blue

Figure 3 illustrates the underlying transitions that occur on addition of a risk factor to the PCE. Every risk stratum is split in two; 0.75 emigrating to a lower risk stratum and 0.25 emigrating to a higher risk stratum. This process is illustrated for select risk strata between 0.06 and .0.09 10-year risk.

**Figure 3.**
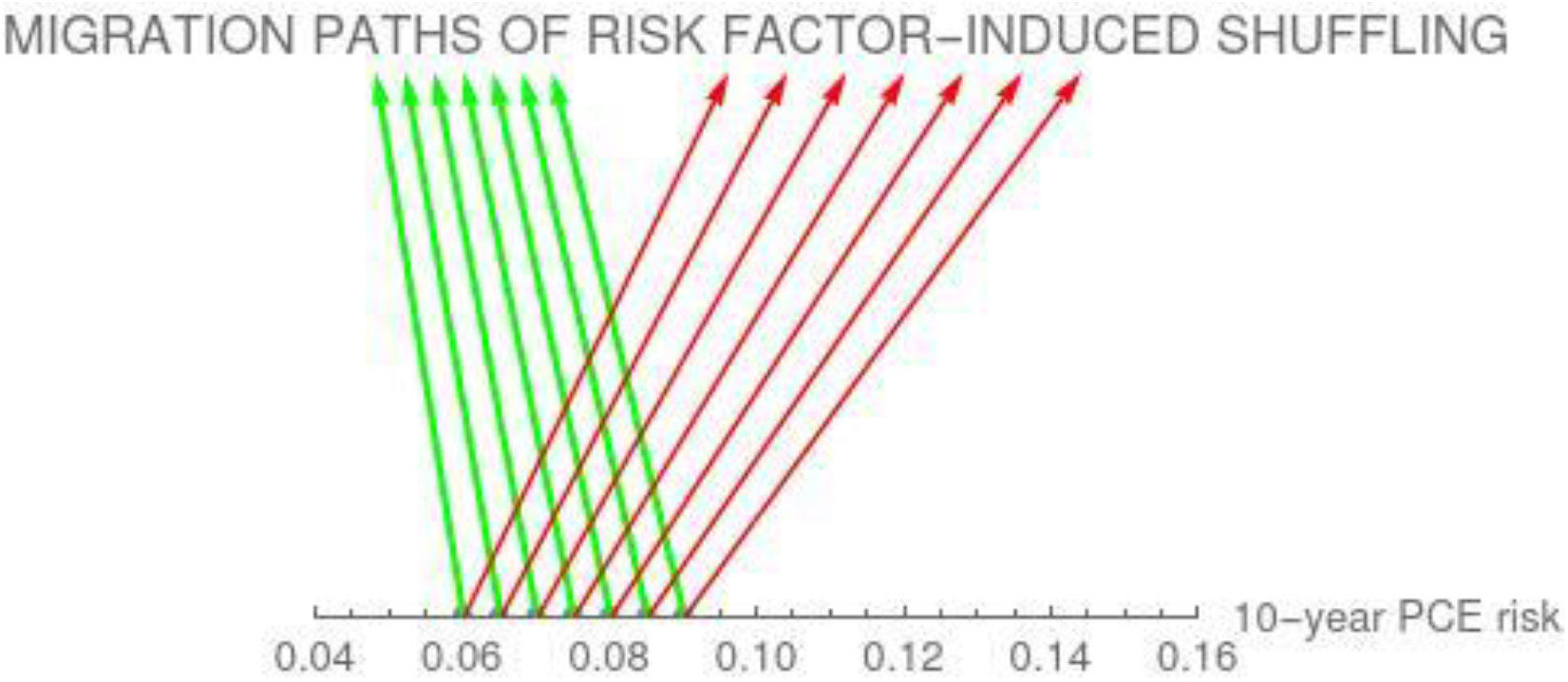
Effect of adding risk factor to PCE 0.75 of each PCE risk stratum migrates to a lower PCE+RF risk stratum (green arrows) and 0.25 to a higher PCE+RF risk stratum (red arrows) on addition of a risk factor

Figure 4 demonstrates that even though the 0.05 PCE risk stratum is completely emptied on addition of a risk factor, the 0.05 PCE+RF risk stratum is refilled by immigration from a lower and a higher risk stratum in the PCE. The PDF densities at 0.05 are 6.03898 for the PCE and 6.02293 for the PCE+RF. The replacement comes from receipt of 0.75 of the 0.0625 PCE risk stratum (0.75*5.07809*1.25) and 0.25 of the 0.03125 PCE risk stratum (0.25*8.07822*0.625

**Figure 4.**
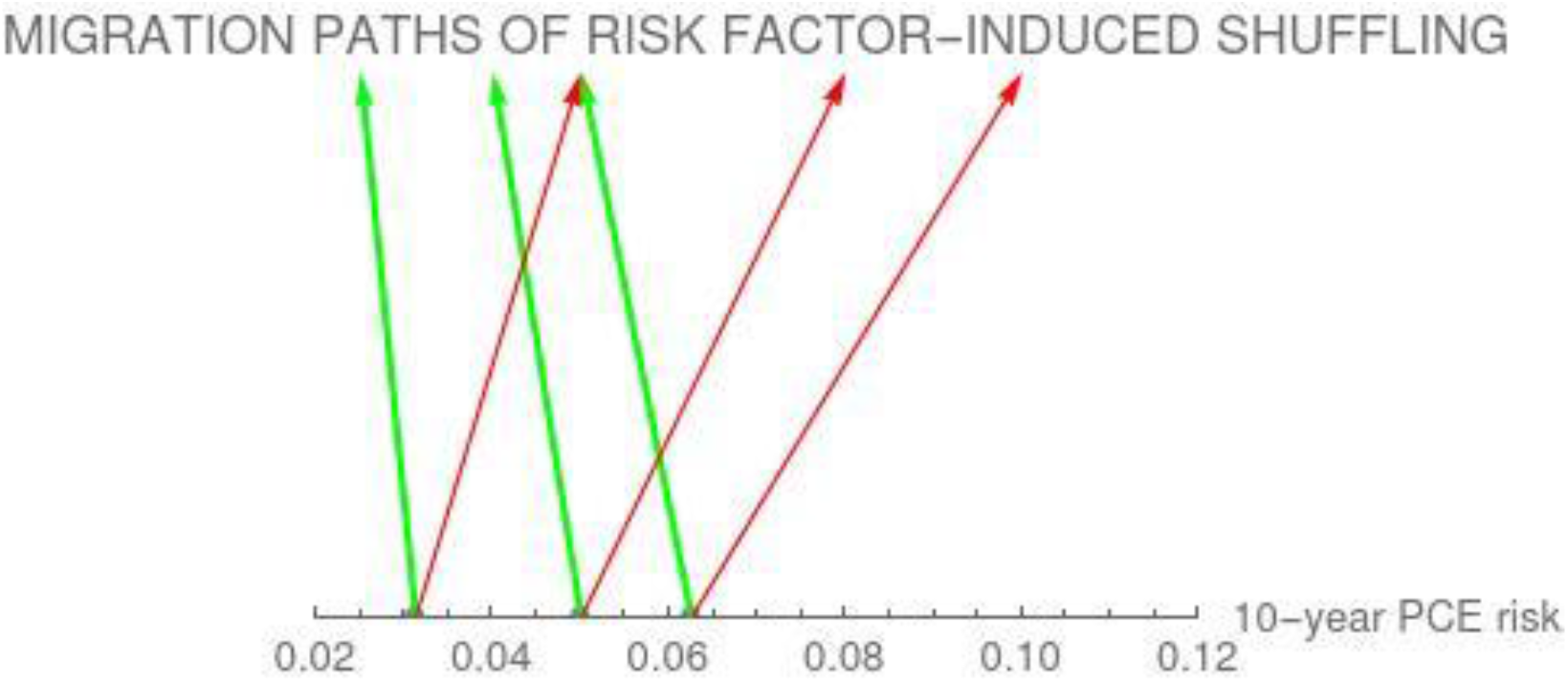
Repopulation of the 0.05 PCE+RF risk stratum on addition of a risk factor 0.75 of each PCE risk stratum migrates to a lower PCE+RF risk stratum (green arrows) and 0.25 to a higher PCE+RF risk stratum (red arrows) on addition of a risk factor, depopulating the 0.05 PCE risk stratum and repopulating the 0.05 PCE+RF stratum

Discrimination is a measure of the separation of the risks of patients/cases and nonpatients/controls. Their risk distribution curves can be calculated from the population risk distributions for the PCE and PCE+RF and are shown in Figure 5. The slightly broader risk distribution on addition of the risk factor leads to slightly greater separation of these two derivative curves. The ROC curves derived from these distributions are shown in Figure 6 and are only slightly different. The ROC curve AUC for the PCE is 0.788361 and for the PCE+RF is 0.800494.

**Figure 5.**
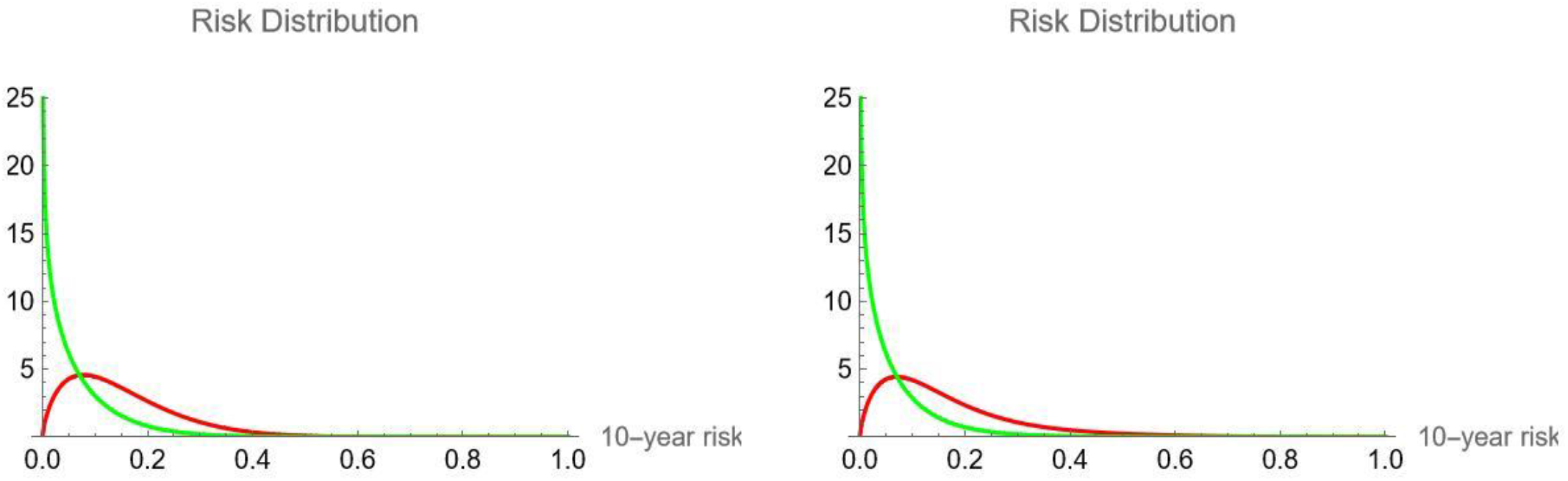
PCE and PCE+RF Risk Distributions of Patients and Nonpatients PCE: Left; PCE+RF Right; Green: Nonpatients; Red: Patients

**Figure 6.**
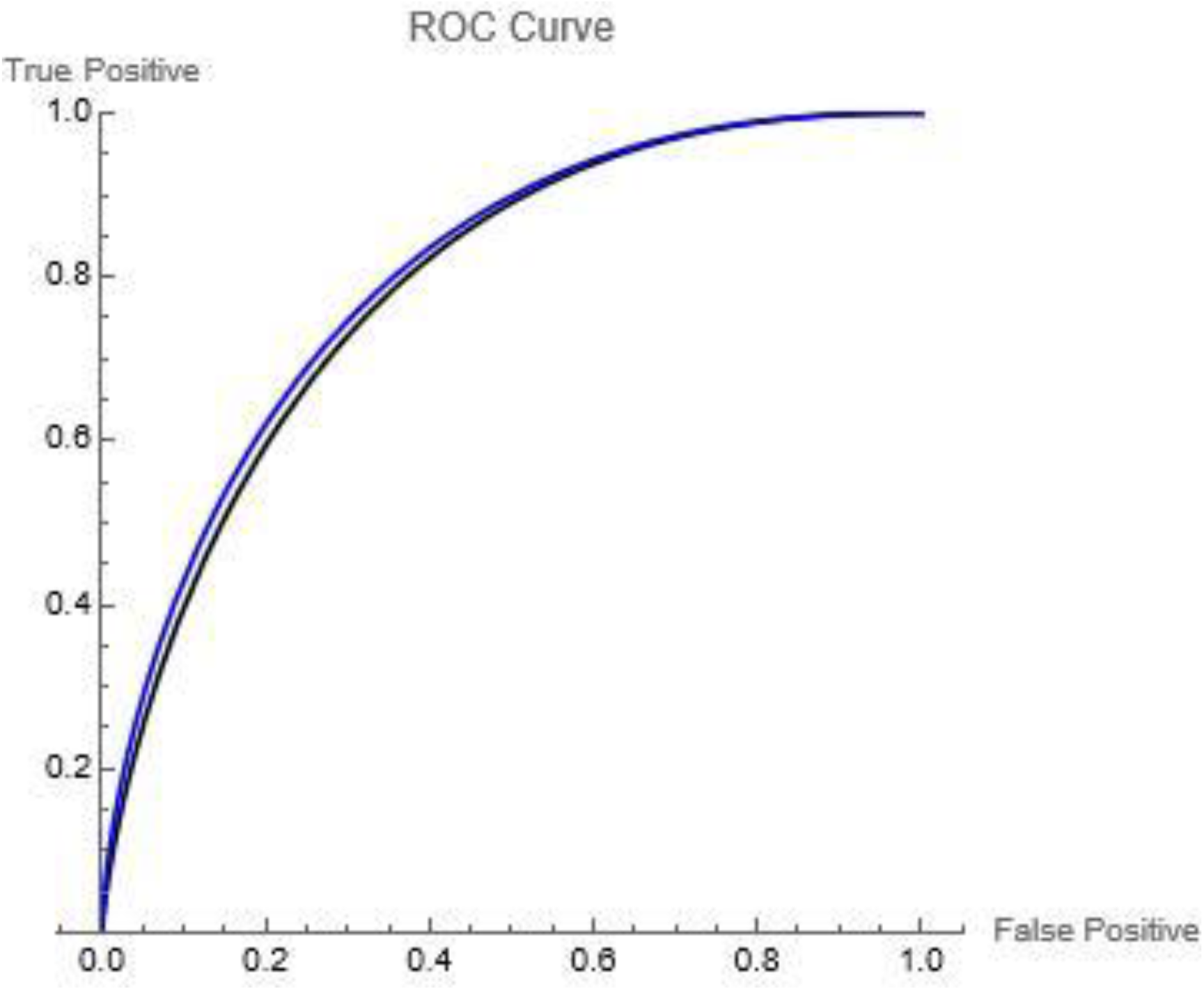
ROC Curves for PCE and PCE+RF PCE: black; PCE+RF: blue

Consistent with the minimal change in the PCE risk distribution on addition of the risk factor, the fraction of the population above a risk of 0.075 in the PCE risk distribution is 0.344262 and in the PCE+RF risk distribution is 0.327146, with the latter being slightly smaller attributed to the skewed distribution with the median below the mean. However, as a result of the shuffling, 0.0367605 of the population is reclassified from below 0.075 in the PCE risk distribution to above 0.075 in the PCE+RF risk distribution, while 0.0538761 of the population is reclassified from above 0.075 in the PCE risk distribution to below 0.075 in the PCE+RF risk distribution. So a total of 0.0906366 of the population is reclassified

## Discussion

Regression models describe the differences in risk between subgroups with different risk factors. These differences are large for a risk factor that doubles risk, as shown in figure 1. On the other hand, discrimination measures describe the differences in risk between subgroups who have (cases or patients) or don’t have (controls or nonpatients) adverse events. The most common approach utilized in the literature is presentation of the ROC curve and its AUC. However the ROC curve is derived from the risk distributions of cases/patients and controls/nonpatients. And these two distributions are derived in turn from the population risk distribution. Thus addition of a risk factor that leads to a broader population risk distribution leads to a greater separation of the risk distributions of cases/patients, which leads to greater separation of the ROC curves and a larger ROC curve AUC or c-index. In contrast to the large difference in risk between those who do and don’t have the risk factor, addition of a risk factor doubling risk to the PCE is associated with minimal changes in the population risk distribution, the separation of the risk distributions for cases/patients and controls/nonpatients, the ROC curves, and the ROC curve AUC. And this is a best case scenario, as most real world risk factors are not completely independent of the components of established models like the PCE.

Although the addition of risk factors triggers universal migration between risk strata, the interweaving of the migration paths results in largely compensatory shuffling of risk assignments. This is the reason large relative risks are required for addition of a risk factor to improve measures of discrimination.

When risk distributions are partitioned into treatment categories, addition of risk factors result in substantial migration across treatment category borders. But again cross-border migrations in opposite directions may largely be balanced.

Since regression analysis and ROC curve AUC analysis provide different information, similar conclusions should not be expected. Of the two, it is the ROC curve AUC that addresses the potential clinical benefit of addition of a new risk factor, as opposed to a statistical benefit. This is because the clinical benefit of selectively assigning preventive measures (treatments or surveillance) based on risk depends on spreading out the risk distribution, which is measured by the ROC curve AUC. Thus, risk factors need to produce an appreciable improvement in the ROC curve AUC to plausibly improve allocation of preventive measures.

Although only a fraction of the population is reclassified on addition of a risk factor, all members of the population are assigned to different risk strata. This results in individuals receiving different risk assignments when models with different risk factors are utilized. This lability, discussed in the philosophy of probability literature as the reference class problem, undermines the narrative that the output from clinical prediction models are individual risks. Their ability to identify subgroups that differ in risk is sufficient justification for their clinical use, but it should be recognized that the risk stratifications they produce are not unique.

## Data Availability

All data produced in the present work are contained in the manuscript

